# Early COVID-related Acute Kidney Injury Recovery May Course with Hydroelectrolytic Disorders in Patients With High Risk of Insensible Fluid Loss

**DOI:** 10.1101/2021.10.20.21265300

**Authors:** Géssica Sabrine Braga Barbosa, Ana Gabriela de Jesus Torres de Melo, Rayra Gomes Ribeiro, Daniela del Pilar Via Reque Cortes, Carla Paulina Sandoval Cabrera, Rubens Santos Andrade Filho, Guilherme Tamborra Pantaroto, Bruno de Castro Paul Schultze, Gilberto Alvarenga Paula, Camila Eleuterio Rodrigues

## Abstract

**Background:** Acute Kidney Injury (AKI) may occur in more than 30% of COVID hospitalized patients, and renal recovery is poorly described.

**Aim:** We aimed to evaluate the renal short-term recovery profile of COVID-related AKI (COV+) compared to COVID-unrelated AKI (COV-).

**Design:** case-control retrospective single-center study

**Methods:** All patients admitted to the Hospital das Clínicas, University of São Paulo, who recovered AKI from April to June of 2020 (COV+, n=98) and from August to October of 2019 (COV-, n=50) were analyzed. Recovery was defined by spontaneous serum creatinine drop or withdrawal of dialysis. Serum electrolytes were analyzed during the first five days of recovery.

**Results:** Among 333 COV+ patients, 98 recovered from AKI (29.4%), while 50 of 177 COV-patients recovered (28.2%). The COV- group presented higher prevalence of chronic morbidities, while the COV+ group had a worse acute clinical course requiring vasoactive drugs (VAD), mechanical ventilation (MV) and dialysis. COVID-19 diagnosis was associated with need of mecxhanical ventilation, dialysis, presence of fever, and higher use of any diuretic drug during first days of recovery. The presence of fever and mechanical ventilation were the predictors associated with intravascular volume depletion surrogates (daily progressive rising in sodium levels and elevation in serum urea: creatinine ratio). Neither COVID-19 nor diuretics use seem to be independent risk factors for this.

**Conclusions:** Intravascular volume depletion surrogates are more common in short-term AKI-recovery of patients presenting fever and mechanical ventilation, commons features in SARS-CoV2 infection.

## Introduction

Since December 2019, a novel virus called ‘severe acute respiratory syndrome coronavirus 2’ (SARS CoV-2) has caused an international outbreak of respiratory illness described as COVID-19. At October 2021, up to 230 million COVID-19 cases, including almost 4.8 million deaths were reported by The World Health Organization (1).

Initial reports indicated that rates of acute kidney injury (AKI) related to COVID-19 were negligible (2). However, growing evidence has demonstrated that AKI is in fact frequent among patients with COVID-19, particularly within patients in the intensive care unit (ICU) (2). Available evidence suggests that it likely affects >20% of hospitalized patients and >50% of patients in the ICU (2).

The pathophysiology of COVID related AKI currently relies on several etiologies starting from unspecific mechanisms such as hypovolemia, nephrotoxic drugs, high positive end expiratory pressure, right heart failure, passing by a direct viral injury, imbalanced Renin-Angiotensin-Aldosterone System (RAAS) activation, elevation of pro-inflammatory cytokines elicited by the viral infection, and procoagulant state (2). Volume depletion at admission might be a common trigger for AKI, as patients with COVID-19 typically present with fever and pre-hospital fluid resuscitation is rarely performed as an attempt to avoid fluid overload and pulmonary edema (3).

The use of diuretics, restriction of volume and renal replacement therapy were strategies intensely used aiming to accomplish negative fluid balance, primarily to improve ventilatory performance (3). Howbeit electrolyte disorders like hypernatremia, metabolic alkalosis and hypovolemia are adverse events associated with negative fluid balance strategies (4, 5). Metabolic alkalosis itself can reduce neural respiratory drive and lead to lower minute-volume contributing to the observed mechanical ventilation weaning difficulties (6). These events can delay AKI recovery during the hospitalization period.

Our study aimed to evaluate the recovery profile of COVID-related AKI (COV+) compared to COVID-unrelated AKI (COV-), depicting clinical characteristics and laboratorial findings, especially fluid balance, electrolyte and acid-base disorders.

## Methods

### Study design and population

We performed a case-control retrospective single-center study in Hospital das Clínicas, University of São Paulo, a large tertiary care university hospital that was solely dedicated to COVID-patient care during the pandemic in 2020.

AKI was determined according to Kidney Disease Improving Global Outcomes 2012 (KDIGO) criteria (7), and recovery of renal function was defined by two consecutive spontaneous serum creatinine drop or definite withdrawal of dialysis. Deceased patients during the illness were considered as non-recovering AKI.

We screened all hospitalized patients with positive COVID test who were evaluated by the AKI nephrology team, from April to June 2020. COVID-19 test included either serologic testing or polymerase chain reaction (PCR) in naso-oropharyngeal swab or tracheal aspirate. Patients with COVID-related AKI who presented recovery of renal function (COV+) were included as cases. The control group included patients hospitalized and followed by nephrology group because of AKI from August to October of 2019, and who presented recovery of renal function (COV-).

Exclusion criteria were: CKD on dialysis, patients without AKI recovery, and those with insufficient data. Only first AKI episode was included in the analysis. Kidney transplant recipients and pregnant woman were not considered for the purpose of this study.

Clinical and laboratorial data were collected at time of AKI diagnosis by reviewing medical records. Biochemical parameters were analyzed in blood during the first five days of recovery of renal function. We used two laboratorial variables as indicators of possible volume depletion in patients studied: concentration of serum sodium, and serum urea:creatinine ratio. We decided not to include other potential surrogates of volume depletion, as serum chloride or gasometric analysis, due to low sequential measures during follow-up. This is also the main reason for choosing surrogate measures instead of the observed fluid balance, as many patients were out of ICU setting and did not present a reliable sequential fluid balance routine.

Diuretic use during the first five days of recovery was assessed. In order to compare doses among patients using different diuretic drugs, we defined that a dose of 20mg intravenous furosemide would be equivalent to 25mg oral hydrochlorothiazide or to 50mg oral spironolactone. Mean diuretic dose along the 5-day recovery period was calculated as the total dose along 5 first days after recovery divided by 5 days.

The primary outcomes were rising values of surrogate markers of intravascular volume depletion, namely serum sodium concentration and serum urea: creatinine ratio, during the first 5 days of AKI recovery in COV+ and COV-patients.

### Ethical Aspects

The study was approved by the local institutional review board (Reference no. 4.112.403), and it is in accordance with the Strengthening the Reporting of Observational Studies in Epidemiology statement (8).

### Statistical Analysis

Categorical variables were presented as number of individuals and percentage, whereas continuous variables were presented as mean ± standard deviation (SD) or median [interquartile range (IQR)] accordingly to the distribution. Student t test or Mann-Whitney test were used to compare groups, and categorical variables were analyzed using Fisher tests. We considered p values <0.05 as statistically significant.

We considered that our surrogate intravascular volume depletion markers (serum sodium concentration and serum urea: creatinine ratio) were our response variables.

We considered the interactions among variables to generate Generalized Estimating Equations (GEE) models based on the day following kidney recovery and clinically relevant variables, as diuretic use, dialysis, mechanical ventilation, presence of fever and COVID-19 diagnosis. We included presence of diarrhea as a variable only in the model that considered serum urea: creatinine ratio, but not for serum sodium concentration model, as depending on the cause and source, diarrhea may increase or decrease sodium levels. Three patients (2 in COV+ group and 1 in COV-group) did not have any information about fever status, being excluded for modeling analysis.

GEE models with autoregressive AR (1) correlation structure were chosen based on quasi-likelihood under independence model criterion (QIC), Rotnitzky-Jewells’ Criteria (RJC) and Gosho-Hamada-Yoshimura’s criterion (GHYC). The final variable selection was based on the Wald test for each parameter, maintaining variables with statistically p-value < 0.05.

Statistical Analysis were performed using GraphPad Prism, version 5.0 for Windows, R statistical software, version 4.0.5 and Rstudio, version 1.4.1106 (R Development Core Team, 2020).

## Results

We included in the final analysis a sample consisting of 98 patients in COV+ group and 50 patients in COV-group; more details are provided in the flowchart in Figure 1.

**Figure 1:**
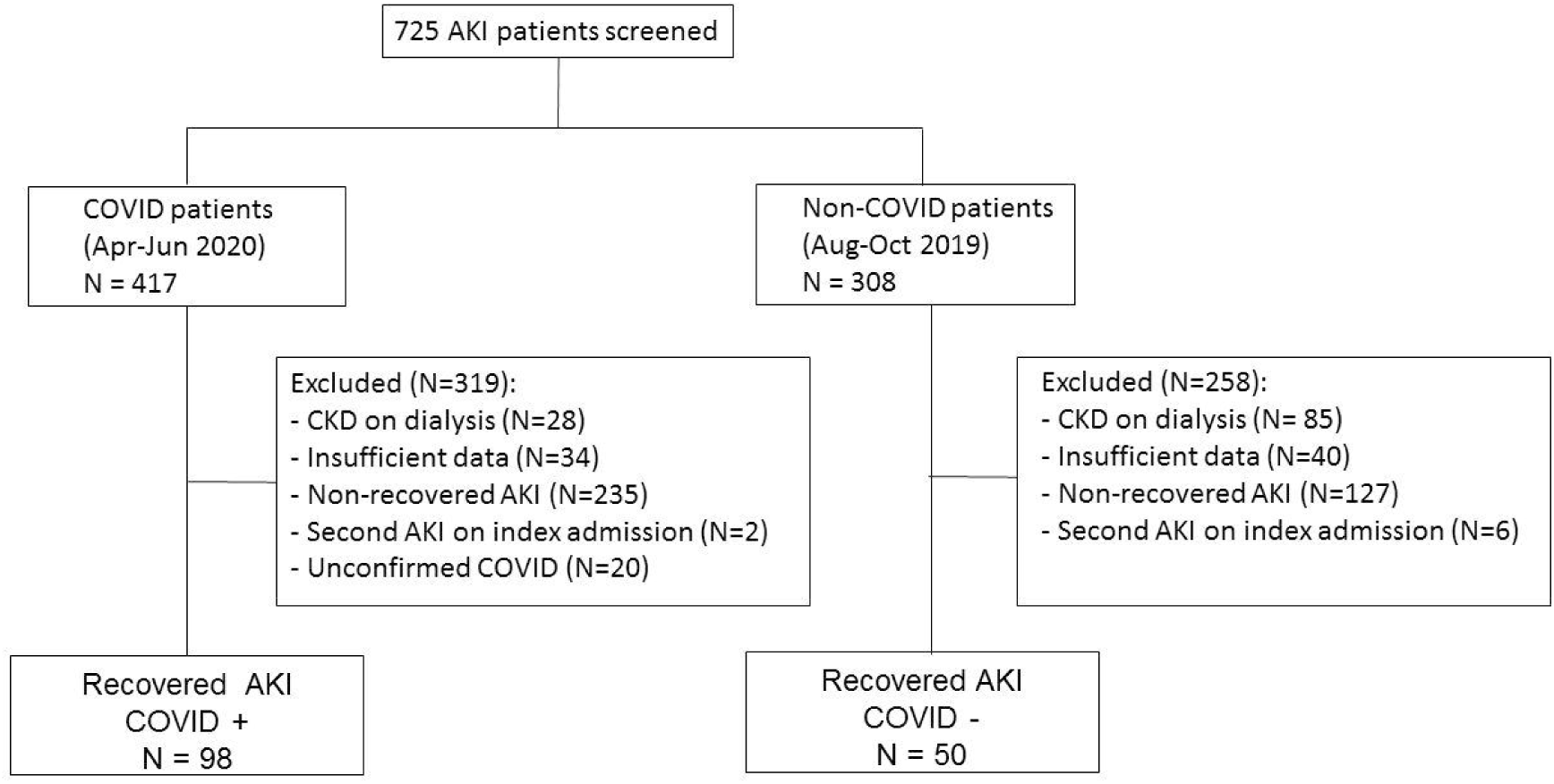
Inclusion criteria population flowchart. AKI: acute kidney injury, COVID: coronavirus disease, CKD: chronic kidney disease.

Among COV+ patients, 98 recovered from AKI (29.4%) and 235 did not, while in COV-group 50 patients recovered (28.2%) and 127 patients did not (p=0.83).

Table 1 shows the demographical and clinical features of all studied patients. There was no difference amongst population age and sex. The COV-group presented higher prevalence of chronic morbidities as chronic kidney disease and peripheral artery disease (p<0.05). Obesity was significantly more common in COV+ patients (p <0.0001).

**Table 1:** Clinical characteristics at time of AKI diagnosis, by COVID-19 status.

|  | COV + (N=98) | COV - (N=50) |
| --- | --- | --- |
| **Age, years, mean ± SD** | 56.3 ± 13.9 | 60.6 ± 13.3 |
| **Male sex, n (%)** | 67 (68.4) | 33 (66.0) |
| **Hypertension, n (%)** | 66 (67.3) | 35 (70.0) |
| **Diabetes, n (%)** | 33 (33.7) | 16 (32.0) |
| **Chronic kidney disease, n (%)\*** | 16 (16.3) | 23 (46.0) |
| **Obesity, n (%)\*** | 40 (40.8) | 1 (2.0) |
| **Peripheral artery disease, n (%)\*** | 1 (1.0) | 6 (12) |
| **Other cardiovascular disease, n (%)** | 12 (12.2) | 10 (20.0) |
| **Vasopressor use, n (%)\*** | 68 (69.4) | 22 (44.0) |
| **Mechanical ventilation, n (%)\*** | 80 (83.7) | 9 (18.0) |
| **AKI KDIGO3, n (%)** | 80 (81.6) | 34 (68.0) |
| **Diuretic use, n (%)\*** | 60 (61.2) | 20 (40.0) |
| **Dialysis, n (%)\*** | 59 (60.2) | 17 (34.0) |
| **Fever, n (%) \*** | 41 (43%) | 11 (22%) |
| **Diarrhea, n (%)** | 12 (12.2%) | 10 (20%) |

\*p < 0.05
SD: standard deviation, AKI: acute kidney injury, KDIGO3: stage 3 AKI accordingly to
Kidney Disease: Improving Global Outcomes criteria

The COV+ group had a worse acute clinical course, presenting higher need of dialysis (p=0.003), vasopressor (p=0.004) and mechanical ventilation (p<0.0001) than COV- group. Any diuretic use (all classes, at any dose) was also higher among COV+ when compared to COV-group (p=0.02), and COVID-19 diagnosis was more associated with presence of fever (p<0.05).

Mean dose of furosemide and hydrochlorotiazide were higher in COV+ patients than in COV-group (Table 2). Only 4 patients in COV+ and 2 patient in COV-group received low-dose spironolactone, resulting in clinically insignificant dosages. Mean daily diuretic dose equivalent to iv furosemide was higher in COV+ group when compared to COV-patients (Table 2).

**Table 2:** Diuretic use in first five AKI-recovery days, by COVID-19 status.

|  | COV + (N=98) | COV - (N=50) |
| --- | --- | --- |
| **Mean iv furosemide dose, mg/day, median [IQR] \*** | 8 [0; 48] | 0 [0; 8] |
| **Mean oral hydrochlorothiazide dose, mg/day, median [IQR] \*** | 2.7 [0; 10.5] | 0 [0; 2.7] |
| **Mean daily diuretic dose, in equivalence to iv furosemide dose, mg/day, median [IQR] \*, †** | 18.1 [0; 55.5] | 0 [0; 18.1] |

\*p < 0.05
iv: intravenous, IQR: interquartile range
Mean diuretic dose was calculated as: the total dose along 5 first days after recovery / 5 days.
† Each 25mg oral hydrochlorothiazide or 50mg oral spironolactone were considered equivalent to 20mg intravenous furosemide

Patients with COVID-19 presented rising serum sodium levels throughout the first five days of recovery, (Figure 2A), and the same pattern occurred with serum urea: creatinine ratio (Figure 2E). Presence of fever (Figure 2B and 2F), mechanical ventilation (Figure 2C and 2G) and use of any diuretic drug (Figure 2D and 2H) were all accompanied by increasing levels of serum sodium and augmentation in serum urea: creatinine ratio during first days of recovery after AKI.

**Figure 2:**
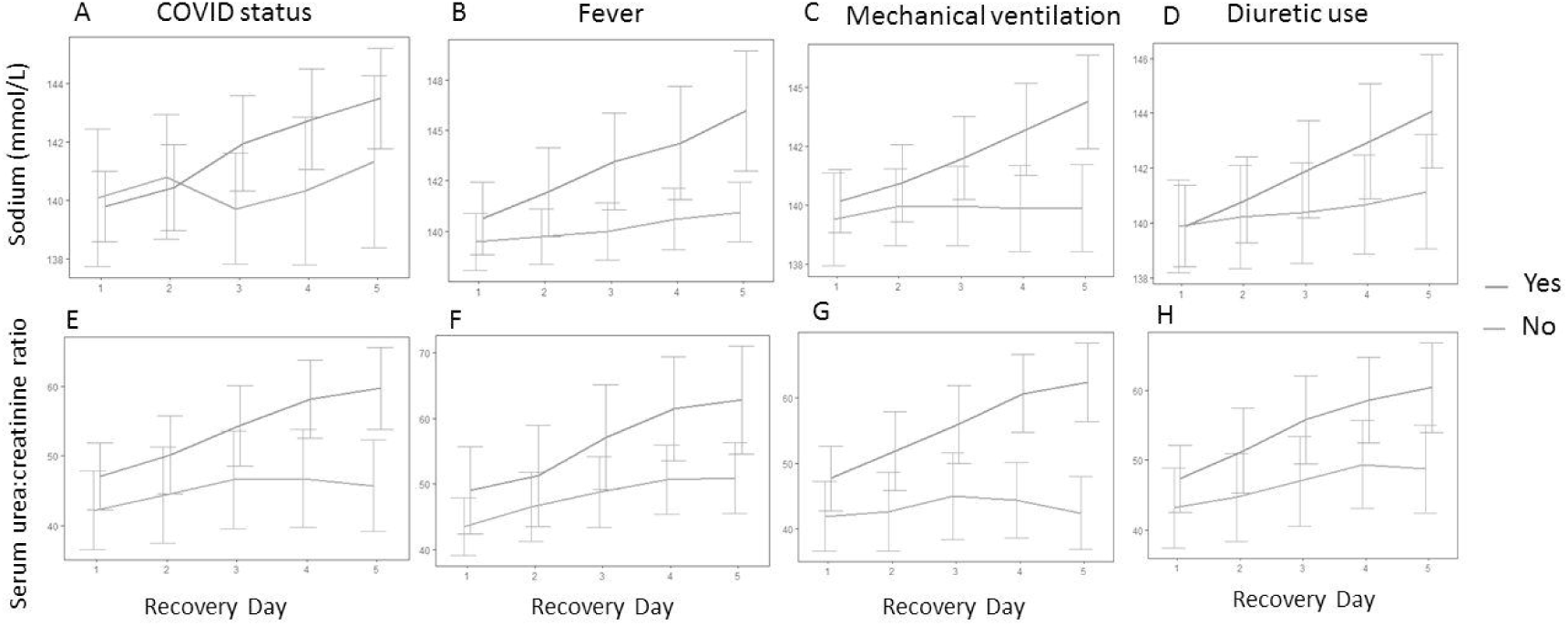
Daily values of serum sodium concentration and serum urea: creatinine ratio on first 5 days after kidney recovery, by COVID-19 status, presence of fever, need of mechanical ventilation and any diuretic use.

GEE with linear normal marginal models and autoregressive AR(1) correlation structure were fitted to explain the serum sodium concentration mean. The mean estimates and the respective confidence intervals, described in Table 3, show that the presence of fever, mechanical ventilation and dialysis were the predictors associated with daily progressive rising in sodium levels. A linear correlation between two successive measures at the same patient was estimated as 0.89 from the GEE normal model. So, for a lag d (=1,2,3,4), the correlation between two measures is estimated as 0.89□.

**Table 3:**
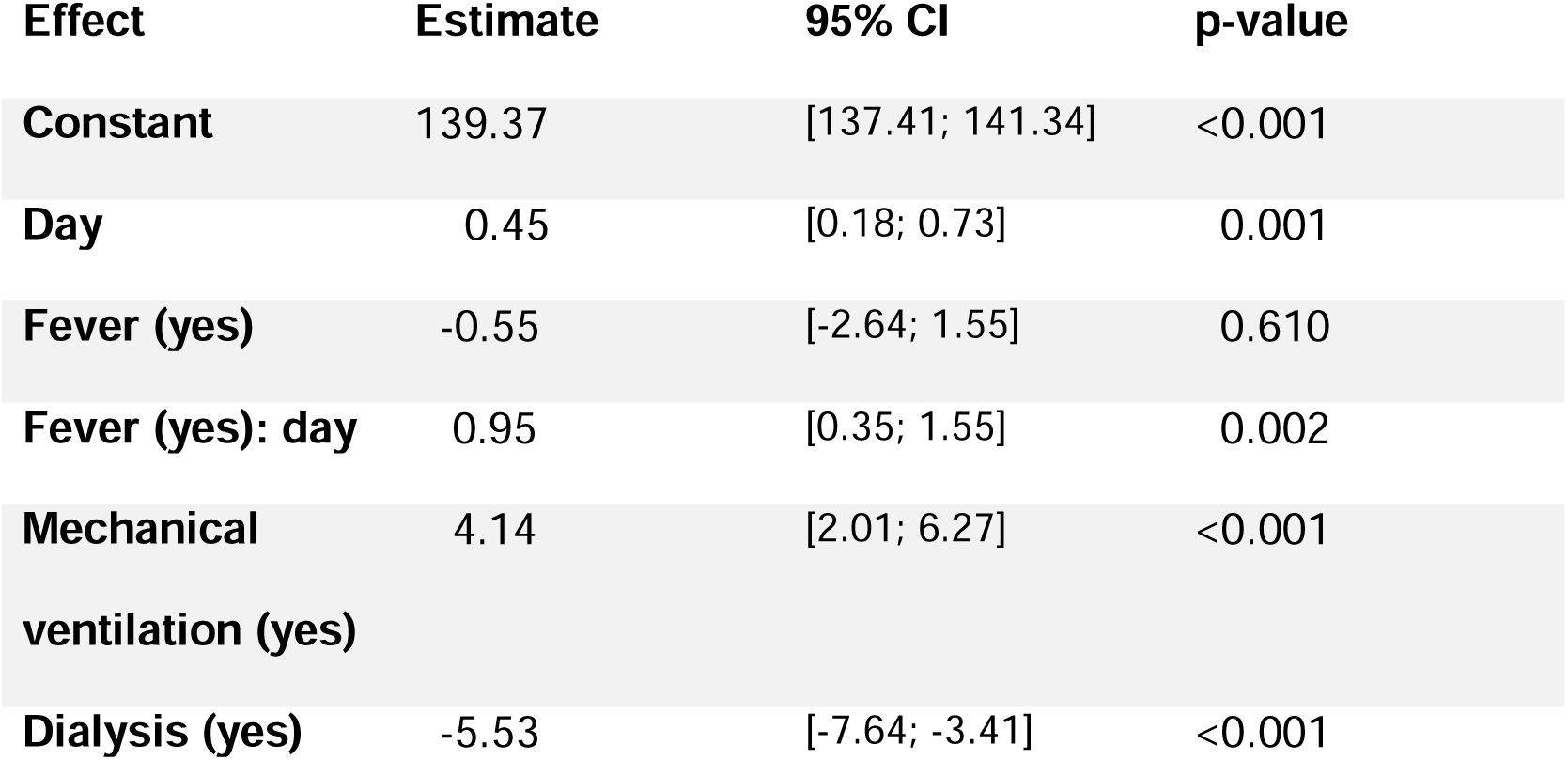
Results from the GEE normal model fitted to explain the serum sodium concentration mean given the predictor variables.

Due to the additive effects of the means in the GEE normal model, various interpretations for each level of the predictor variables may be easily obtained. For example, the patient that does not present fever nor need for dialysis or mechanical ventilation, the mean estimate of serum sodium concentration would be 139.37 + 0.45 = 139.82 mEq/L on the first day of renal recovery, and 139.37 + 2 × 0.45 = 140.27 on the second day of recovery. The presence of fever would increase 0.95 – 0.55 = 0.40 mEq/L in sodium concentration each day (as there is interaction between the variable fever and the day of renal recovery follow-up). Thus, we notice that fever and mechanical ventilation were the main predictors of rising sodium values along renal recovery after AKI.

Similarly, GEE with log-linear gamma marginal models and autoregressive AR(1) correlation structure were fitted to explain the serum urea: creatinine ratio. The mean estimates and the respective confidence intervals are shown in Table 4. The correlation between two successive measures at each patient was estimated as 0.93 from the GEE gamma model. Then, the estimated correlation between two measures of lag d becomes 0.93□.

**Table 4:** Results from the GEE gamma model fitted to explain the serum urea: creatinine ratio given the predictor variables.

| Effect | Estimate | 95% CI | p-value |
| --- | --- | --- | --- |
| **Constant** | 43.27 | [37.74; 49.61] | <0.001 |
| **Day** | 1.01 | [0.99; 1.03] | 0.346 |
| **Mechanical ventilation (yes)** | 1.23 | [1.00; 1.52] | 0.045 |
| **Mechanical ventilation (yes): day** | 1.05 | [1.02; 1.08] | 0.002 |
| **Dialysis (yes)** | 0.78 | [0.67; 0.92] | 0.003 |

Because one has multiplicative effects for the means in the GEE gamma model, the presence of mechanical ventilation would increase the serum urea: creatinine ratio 1.23×1.05 = 1.29 on first day of renal recovery, resulting in a total increase of 29%. On the other hand, the presence of dialysis would decrease the serum urea: creatinine ratio in 22%.

Neither COVID-19 diagnosis nor diuretic use were independent predictors of a rising serum sodium level or of elevation in serum urea: creatinine ratio.

Residual analysis and sensitivity studies (omitted here) were also performed and confirmed the adequacy of the proposed models.

## Discussion

This study was conceived to indicate if COVID-19 could be associated with electrolytic disturbances commonly found in hypovolemic status. We used the increase in serum sodium levels and in serum urea:creatinine ratio as surrogates for intravascular volume depletion in patients admitted to our hospital with and without SARS-CoV2 infection.

We acknowledge that using the rising concentration of serum sodium, and the increase in serum urea:creatinine ratio as surrogates of volume depletion may present some pitfalls, as other conditions may explain these laboratory variations. However, as this was a retrospective study, we could not obtain reliable measurements of direct fluid status variables, as fluid balance, patient weight or sequential chloride levels.

Hypernatremia is the result of a deficit of total body water in relation to total body sodium, what frequently occurs by reduced vascular fluid volume (9). Other causes of hypernatremia may exist, but are extremely uncommon in COVID-19 settings (10). However, volume depletion may be of particular importance in patients with COVID-19, who might present nausea and vomiting, low oral water intake and diarrhea (11). Besides, COVID-19 patients are at higher risk of presenting some factors associated with insensible water loss, as fever and need of mechanical ventilation (12). Fluid overload has not been demonstrated to be a common feature in patients with COVID-19, and pulmonary edema seems to be attributable to other mechanisms, mainly abnormal lung humoral metabolism (13). Despite this, we found that COV+ patients are more prone to receive diuretics than patients recovering from AKI caused by other pathologies.

Previous studies have shown that throughout the course of the disease, COVID-19 may be associated with some electrolyte disturbances, mainly hyponatremia (14, 15). Despite the occurrence of the syndrome of inappropriate antidiuresis (SIAD) has been proposed as a possible mechanism, also present in pneumonias of other etiologies (16, 17), the occurrence of hypovolemic status has been indicated as the major cause of hyponatremia in COVID-19 patients (18). Hyponatremia was not considered a risk factor for in-hospital mortality in COVID-19 patients, except in the case of being caused by hypovolemia (18).

Despite hyponatremia has been reported as the most frequent electrolyte disturbance in COVID-19, in our center, our first impression during clinical rounds was that COVID-19 patients who were presenting renal recovery from AKI apparently presented more hypernatremia than non-COVID-19 patients. They seemed to behaved differently also regarding serum urea levels, as patients with COVID-19 seemed to present higher serum urea: creatinine ratio, resembling a hypovolemic intravascular status that could maybe be attributable to the disease. In fact, we could demonstrate both serum sodium and serum urea: creatinine ratio increase throughout the first days of recovery from AKI. Hypernatremia has previously been described in COVID-19 patients (10, 18, 19), and high sodium levels may be considered a predictor or death in COVID-19 patients (10, 18).

Serum urea concentration has been shown to increase in acute dehydration settings (20, 21), and patients with higher serum urea:creatinine ratios have already been demonstrated to have higher mortality risk than those with lower ratios (22). Indeed, either elevated serum sodium concentrations or increased serum urea:creatinine ratios have been used as measures of volume depletion, and their combined use may be preferred as surrogate markers of hypovolemia (21).

We demonstrated that COVID-19 patients were more exposed to mechanical ventilation, and presented more fever than patients without COVID-19 in this study. In addition, they received higher doses of diuretics. As there was important influence of these multiple factors, COVID-19 status alone was not considered an independent risk factor for hydroelectrolytic disorders.

Instead, mechanical ventilation was an independent factor to hypernatremia and to higher serum urea: creatinine ratio in the recovery period after AKI. Our study also highlighted the importance of fever as a source of insensible water loss, as patients presenting fever were at higher risk of developing rising serum sodium levels throughout the recovery days.

The results show that we should consider occurrence of volume depletion in patients with fever and mechanical ventilation during recovery AKI, associated or not with COVID-19. In this context, the use of diuretics has to be carefully analyzed, due to its potential to get worse electrolytic and metabolic disorders.

We recognize that our study has some important limitations, mainly regarding the low number of patients, the unicentric experience, and the retrospective design. Also, the impossibility to consider the fluid balance of patients certainly impaired the real volume depletion analysis that would be ideally done.

However, to our knowledge this is the first study that recalls some attention to the possibility of hydroelectrolytic disorders in patients who are recovering from AKI in COVID-19 patients, and we believe that this concern needs to exist when caring of patients with potential risk of insensible fluid loss.

## Conclusion

Increase in serum sodium levels and in serum urea:creatinine ratio may be common in short-term AKI-recovery of patients who needs mechanical ventilation or present fever, both commons features in COVID-19 presentation.

## Data Availability

The dataset used and analyzed during this study is available from the corresponding author on reasonable request, and will be found at an international repository (Zenodo) immediately after study publication in a peer-reviewed jounal.

https://zenodo.org/

## Acknowledgements

Some of these data was accepted as an abstract for poster presentation at the American Society of Nephrology (ASN) Kidney Week 2020. It was presented on Thursday, October 22^nd^, in a fully digital meeting.

CER received fees for providing instruction in catheter insertion from Medtronic. All other authors have nothing to disclose.

The dataset used and analyzed during this study is available from the corresponding author on reasonable request, and will be found at international repositories immediately after study publication.

This study was supported by the University of São Paulo School of Medicine Foundation (#HCCOMVIDA initiative for COVID-related Research at the University of São Paulo School of Medicine Hospital das Clínicas).

## Notes

### Funding Statement

This study was supported by the University of Sao Paulo School of Medicine Foundation (#HCCOMVIDA initiative for COVID-related Research at the University of Sao Paulo School of Medicine Hospital das Clinicas).
No other funding was received.

### Author Declarations

The study was approved by the Ethics committee in the Hospital das Clinicas, University of Sao Paulo School of Medicine: Comissao de Etica para Analise de Projetos de Pesquisa - CAPPesq (Reference no. 4.112.403)

